# Metagenomic and single-cell RNA-seq survey of the H. pylori-infected stomach in asymptomatic individuals

**DOI:** 10.1101/2021.12.04.21267139

**Authors:** Chiara Sorini, Kumar P. Tripathi, Shengru Wu, Shawn M. Higdon, Jing Wang, Liqin Cheng, Sanghita Banerjee, Annika Reinhardt, Taras Kreslavsky, Anders Thorell, Lars Engstrand, Juan Du, Eduardo J. Villablanca

**Affiliations:** Division of Immunology and Allergy, Department of Medicine Solna, Karolinska Institutet and University Hospital, Stockholm, Sweden; Center of Molecular Medicine, Stockholm, Sweden; Department of Microbiology, Tumor and Cell Biology, Centre for Translational Microbiome Research (CTMR), Karolinska Institutet, Stockholm, Sweden; Ersta Hospital, Stockholm, Sweden

## Abstract

**Objective:** *Helicobacter pylori* colonization of the gastric niche can persist for years in asymptomatic individuals. Although latent *H. pylori* infection can progress to cancer, a detailed survey of the microbiome and immune composition in the chronically infected stomach is still lacking.

**Design:** We collected human gastric tissues and performed metagenomic sequencing, single-cell RNA sequencing (scRNA-seq), flow cytometry and fluorescent microscopy to deeply characterize the host-microbiota environment in *H. pylori-*infected (HPI) stomachs.

**Results:** HPI asymptomatic individuals showed dramatic changes in the composition of gastric microbiome and immune cells compared to non-infected individuals. With metagenomic data, we also demonstrated antibiotic resistant genes, enzymes and pathway alterations related to metabolism and immune response. scRNA-seq and flow cytometry data revealed that in contrast to murine stomachs, ILC2 are virtually absent in the human gastric mucosa, whereas ILC3 are the dominant population in asymptomatic HPI individuals. Specifically, NKp44^+^ ILC3s were highly increased in the gastric mucosa of asymptomatic HPI individuals, and their proportions correlated with the abundance of selected microbial taxa found to be enriched in the infected mucosa. In addition, CD11c^+^ myeloid cells, activated CD4 T cells and B cells were expanded in HPI individuals. In HPI individuals, B cells acquired an activated phenotype and progressed into a highly proliferating germinal center stage and plasmablast maturation, which correlated with the presence of tertiary lymphoid structures within the gastric lamina propria.

**Conclusion:** Our study provides a comprehensive atlas of the gastric mucosa-associated microbiome and immune cell landscape when comparing asymptomatic HPI and uninfected individuals.

**SIGNIFICANCE OF THE STUDY:** What is already known on this subject?

- Previous studies on the gastric microbiome were performed via 16S rRNA gene sequencing.
- Acute *Helicobacter* spp. infection in murine models and symptomatic *H. pylori*-driven pathology in humans result in remodeling of the stomach immune cell compartment.
- ILC2 is the dominant ILC population in the murine stomach.

What are the new findings?

- We described the effect of chronic asymptomatic *H. pylori* infection on the gastric microbiome via whole-genome sequencing.
- Single cell census of the gastric mucosa reveals ILC3 to be the dominant ILC population in the human stomach, whereas ILC2 were virtually absent.
- scRNA-seq reveals the gastric immune cell programs in asymptomatic *H. pylori*-infected individuals, which is characterized by the formation of tertiary lymphoid structures.

How might it impact on clinical practice in the foreseeable future?

- Whole genome sequencing of uninfected and *H. pylori*-infected gastric mucosa bolsters collective knowledge of stomach physiology with respect to the gastric microbiome and microbiota function.
- We present a comprehensive immune cellular landscape of the human stomach, which will be a valuable resource to interrogate pathology of gastric diseases.

## INTRODUCTION

*Helicobacter pylori* chronically infects the gastric epithelium of approximately 50% of the world’s population ^1^. Symptoms associated with chronic inflammation caused by *H. pylori* include loss of appetite, abdominal pain, and unintentional weight loss. Moreover, chronic inflammation induced by *H. pylori* is associated to an increased risk of developing severe gastric pathologies, including peptic ulcer and cancer. Next-generation sequencing technology has enabled characterization of the gastric microbiome, which is dominated by the Actinobacteria, Bacteroidetes, Firmicutes and Proteobacteria in homeostatic conditions ^2^. The bacterial ecosystem of the gastric niche is significantly affected by *H. pylori* infection ^3^, which may contribute to the broad spectrum of clinical manifestations in chronically infected patients ^4–6^. However, how *H. pylori* infection influences the composition and function of the overall gastric microbiome is yet to be investigated.

*H. pylori* can prompt gastric epithelial cells to secrete cytokines and chemokines, which eventually recruit innate and adaptive immune cell types ^7^. Among immune cells/cytokines associated with active protection against infection, IL-17A levels inversely correlated with *H. pylori* load in the gastric mucosa, suggesting a role of IL-17-secreting cells in eradicating acute *H. pylori* infection ^8, 9^. However, the presence of T helper 17 cells has been linked to more severe gastric inflammation and tumor progression in adult patients ^10^. A recent study on mice with acute *H. pylori* infection linked B cell numbers and *H. pylori*-specific IgA responses in the gastric mucosa to an expansion of type 2 innate lymphoid cells (ILC2) ^11^. The presence of these and other immune cell types in the human stomach, their crosstalk with the human gastric microbiome, and their contributions to human pathology remain as topics of debate.

Importantly, 80-90 % of infected individuals develop long-term infection and gastritis remains asymptomatic ^12^; however, whether the changes observed in chronic inflamed *H. pylori*-infected (HPI) individuals are also present in those who are asymptotic is currently unknown. Here, we characterized the gastric microbial microenvironment and immune cell types that accumulate in the gastric mucosa of asymptomatic HPI individuals.

## RESULTS

### Composition of the gastric microbiome is severely altered during *H. pylori* infection

Despite the known gastric microbiome changes related to *H. pylori* infection, it is still unclear if the infection affects bacterial species in different gastric regions ^5, 6, 13^. We collected samples from fundus and antrum of five patients undergoing gastric sleeve surgery, each of which lacked gastritis-related symptoms. *H. pylori* quick test and *H. pylori* culture results showed that two patients were *H. pylori*-infected at the time of sample collection. Mucosal swab samples then underwent shotgun whole genome sequencing to functionally characterize the microbiome and describe its bacterial composition (figure 1A).

**Figure 1.**
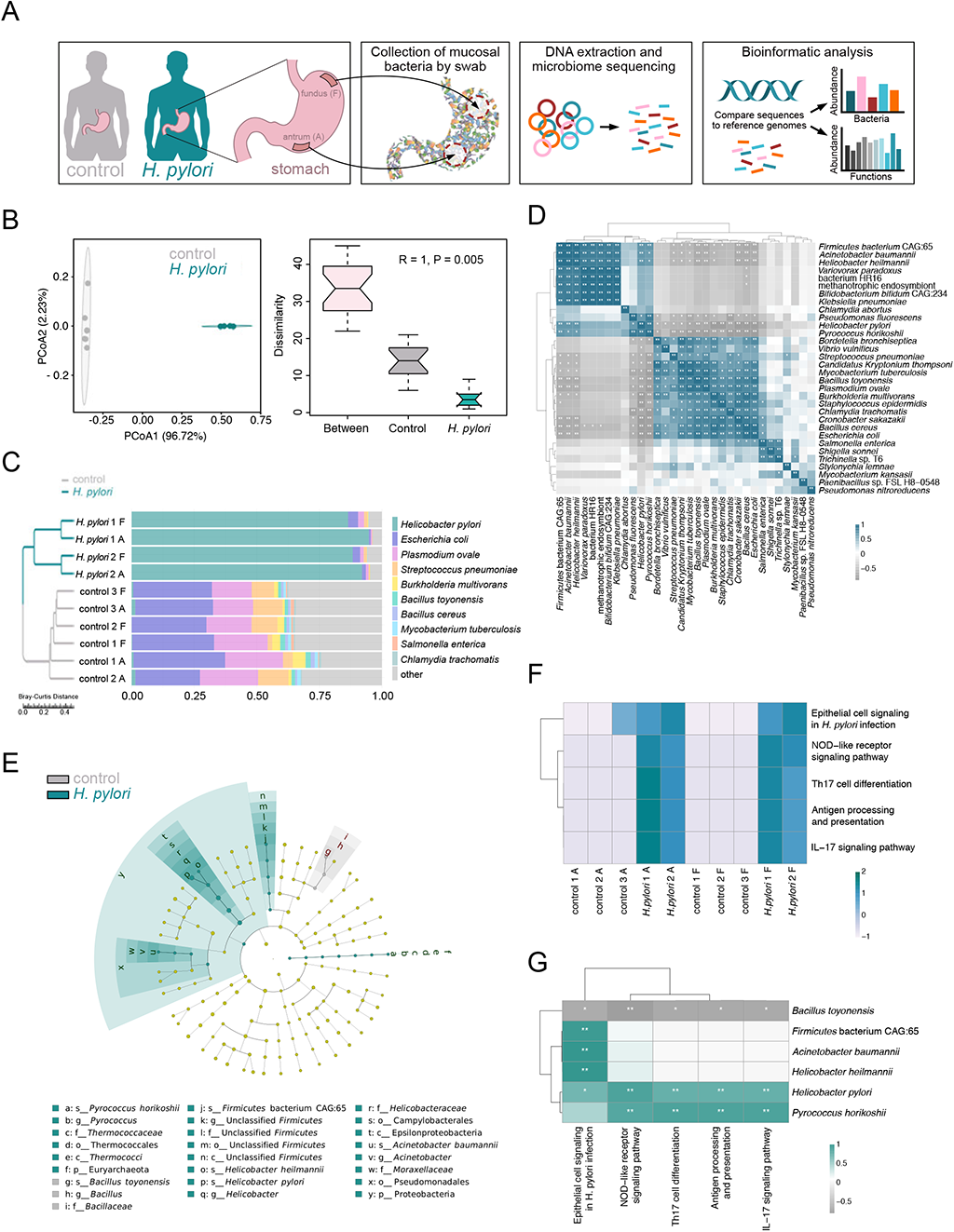
Composition and immune related functions of the gastric microbiome are altered in HPI tissues. **a.** Schematic representation of experimental workflow for detecting the gastric microbiome from sample collection, through DNA extraction to metagenomic sequencing and analysis. **b.** The principal coordinates analysis (PCoA) with identified bacterial species based on Bray-Curtis distance. The analysis of similarities (Anosim) showing a significant difference in the microbial diversity between the HPI and uninfected tissues. **c.** The ten most abundant bacterial species and sample cluster of the HPI and uninfected tissues based on Bray-Curtis distance. F: Fundus, A: Antrum. **d.** The Spearman correlation among the identified most abundant species. \**p* < 0.05, \*\**p* < 0.01, \*\*\**p* < 0.001. **e.** Differential bacteria were identified between the HPI and uninfected tissues using linear discriminant analysis (LDA) Effect Size (LEfSe) analysis (LDA>4). **f.** Significantly increased microbial functional pathways that related to immune function were identified in HPI tissues. The differential metagenome functions were obtained using Metastats analysis with *q* value < 0.05. **g.** The spearman correlation analysis based on the identified differential species and immune related KEGG pathways between the *H. pylori*-infected and uninfected tissues. \**p* < 0.05, \*\**p* < 0.01, \*\*\**p* < 0.001

The gastric microbiome composition was significantly altered in *H. pylori*-infected (HPI) compared to non-infected individuals (figure 1B), and the difference between the two groups was bigger than the difference within each group (pANOSIM = 0.005). The metagenomic sequencing data confirmed that infected tissues were heavily dominated by *H. pylori,* while the uninfected samples had major proportions of *Escherichia. coli*, *Plasmodium ovale,* and *Streptococcus pneumoniae* (figure 1C). Concurrent results were observed at the phylum level (suppl. figure 1A). Correlation analysis illustrated potential mutualistic interactions between *H. pylori* and other species of the gastric microbiome. *Helicobacter heilmannii* and *Acinetobacter baumannii*, which both had increased relative abundances in HPI tissues, were positively correlated with a high relative abundance of *H. pylori,* and inversely correlated with the high relative abundance of *Bacillus toyonensis, which* was significantly higher in uninfected tissues (figure 1D, E). In addition, *E. coli*, *Bacillus cereus* and *Cronobacter sakazakii* had positive relative abundance correlations with *B. toyonensis*, but were all inversely correlated with that of *H. pylori.* No heterogeneity in the microbiome was identified between the fundus and antrum samples (suppl. figure 1B). These data suggest the potential relationship of different microbes with *H. pylori* and the establishment of altered microbiota composition in HPI asymptomatic individuals.

### Gastric microbiome is functionally linked to immune response

We sought to investigate whether *H. pylori*-induced dysbiosis affects the host immune response. Assignment of the shotgun metagenomic reads to the Kyoto Encyclopedia of Genes and Genomes (KEGG) pathway and Carbohydrate-Active enzymes (CAZy) databases ^14, 15^ showed the presence of microbial functions that were evident and distinct between HPI and uninfected tissues (suppl. figure 1C). Microbial function pathways, especially those pertaining to metabolic processes, immune system and infection, were dramatically increased in HPI tissues (suppl. figure 1D). Genes conferring glycosyltransferases (GT) from families GT8, GT9, GT25, and a carbohydrate binding module (CBM) from CBM50 were significantly increased in HPI tissues (suppl. figure 1E), with most enzymes being involved in pathways such as biosynthesis and metabolism of lipopolysaccharides (LPS) and glycans (suppl. figure 1F) ^16^. Furthermore, all samples from HPI tissues contained the hp1181 gene, which serves as an important antibiotic resistance gene associated with the active efflux and multidrug-resistant phenotype (suppl. figure 1G) ^17, 18^. Analysis of immune related pathways showed a significant increase in epithelial cell signaling, antigen processing and presentation, Th17 cell differentiation and IL-17 signaling, and NOD-like receptor signaling among HPI tissues (figure 1F). These pathways are widely involved in both B cell and T cell functions. Evaluating correlations between relative abundances for sample microbiota composition and functional pathways also indicated that increased *H. pylori* and *Pyrococcus horikoshii* values along with decreased *B. toyonensis* population sizes were linked to an elevated presence of genes conferring the aforementioned immune pathways (figure 1G). Collectively, these findings suggest the potential for gastric microbiome and immune compartment interplay with a leading role of *H. pylori*.

### scRNA-seq resolves the lymphoid and myeloid gastric immune compartment

To test the hypothesis that dysbiotic gastric microbiota affect the gastric mucosal immune compartment, we characterized the immune cell composition of healthy and HPI individuals using scRNA-seq in an unbiased fashion (figure 2A). Cells from the fundus and antrum regions of the HPI and uninfected gastric tissues were differentially barcoded with oligonucleotide-conjugated antibodies and sorted by fluorescence-activated cell sorting (FACS) into three main cell populations: Lin^-^CD127^+^ cells (innate lymphoid cells, ILC), Lin^+^ cells (T cells) and Lin^-^CD127^-^HLA-DR^+^ cells (containing myeloid and B cells and henceforth referred to as HLA-DR^+^) (figure 2A). Integration and merging of all datasets resulted in 22033 high-quality cells (19774 genes expressed in total), which were manually annotated based on signature genes (figure 2B-E). Five clusters represented different B cell activation stages (*CD19*^+^*MS4A1*^+^) and plasmablasts (*PRDM1*^+^*CD38*^+^), three clusters represented classical T cells (*CD3*^+^*CD4*^+^ or *CD3*^+^*CD8*^+^), one cluster represented ILC (highest average expression of *IL7R* and lack of *CD3),* and two clusters were annotated as myeloid cells (*HLA-DRA* and *ITGAX)* (figure 2D). Annotation of T and B cells was supported by expression of T cell receptor and immunoglobulin-encoding genes respectively (figure 2E), and all identified populations belonged to the expected sorted fractions (figure 2F). Analysis of the fundus and antrum-associated barcodes revealed no clear separation of cell types based on the stomach region (figure 2G and suppl. figure 2B), which is in line with the non-regionalized microbiome composition previously observed (suppl. figure 1B). Altogether, scRNA-seq enabled the visualization of innate and adaptive immune cells in the gastric mucosa.

**Figure 2.**
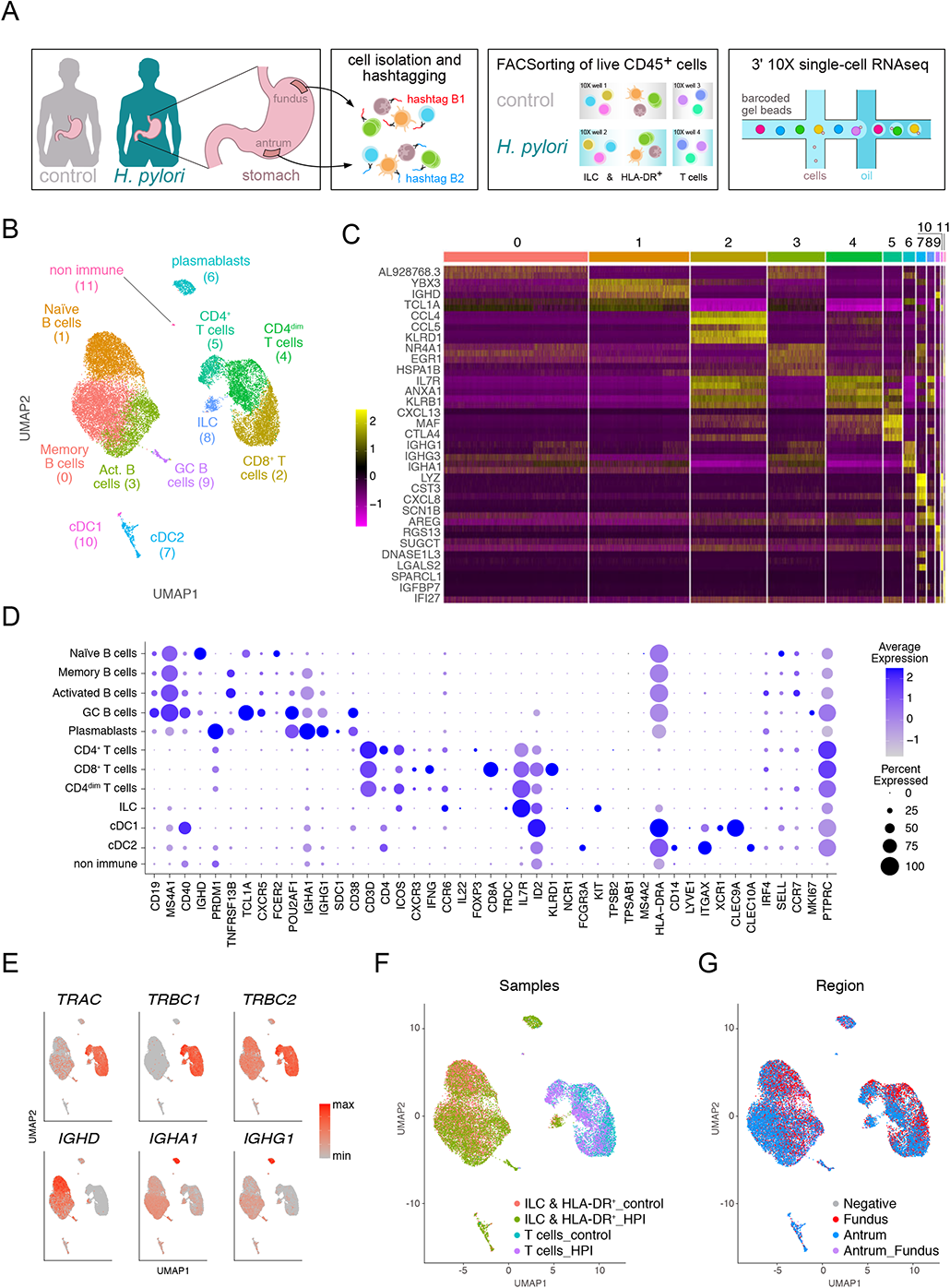
scRNA-seq resolves the lymphoid and myeloid gastric immune compartment. **a.** Schematic representation of the experimental workflow leading from the isolation of gastric immune cells, through hash tagging and sorting to 3’ single-cell RNA isolation and sequencing. **b.** Uniform Manifold Approximation and Projection (UMAP) showing unbiased clustering analysis of all sequenced cells. **c.** Heatmap of top differentially expressed genes between clusters shown in panel **b**. **d.** Dotplot showing expression of selected cell markers that were used to annotate clusters in panel **b**. **e.** UMAP plots showing expression of selected T cell receptor and immunoglobulin genes. **f.** UMAP visualization of all sequenced cells, color-coded based their origin from each sorted fraction. **g.** UMAP visualization of all sequenced cells, color-coded based on fundus and antrum regions. Data represent a pool of cells from the gastric mucosa of three *H. pylori*-infected and six uninfected individuals.

### Gastric innate cell composition in control and HPI individuals

To further characterize the immune cell compartment in the gastric mucosa, we first focused on the ILC & HLA-DR^+^ samples (figure 2A and suppl. figure 2A). After *in silico* exclusion of B cells and plasmablasts, the enriched myeloid and innate lymphoid cells yielded 5 distinct clusters (figure 3A, B). Three clusters of myeloid cells were characterized by high *HLA-DRB1* and *ITGAX* expression. Monocytes (cluster 2) were defined by the expression of *FCGR3A* (CD16) and a non-classical gene expression profile (*CDKN1C*, *LILRB2*) ^19^, as well as *FPR2*, a marker related to chemotaxis to inflamed tissues ^20, 21^ (figure 3C and suppl. figure 3A). cDC1 (cluster 4) was identified by *CLEC9A*, *IRF8, XCR1*, *BATF3* expression, while cDC2 expressed, among other genes, *CD1C*, *CD14* and *SIRPA* (figure 3A-C and suppl. figure 3A). Chronic *H. pylori* infection affected cell proportions with cDC2 being the predominant population in the myeloid compartment (figure 3D). cDC2 from HPI individuals expressed higher *GPR183* (encoding EBI2) levels compared to cDC2 from uninfected gastric tissues, suggesting the formation of ectopic lymphoid follicles (figure 3E) ^22^. Cluster 0 was annotated as innate lymphoid cells (ILCs), mainly defined by their high expression of *IL7R* and *KLRB1* (CD161) (figure 3A, B). Among the most differentially expressed genes in the ILC cluster compared to the rest, *SPRY1*, *XCL1*, *LTB*, *CD2* and *KIT* (suppl. figure 3A) suggest that these cells might contain ILC3 ^23, 24^.

**Figure 3.**
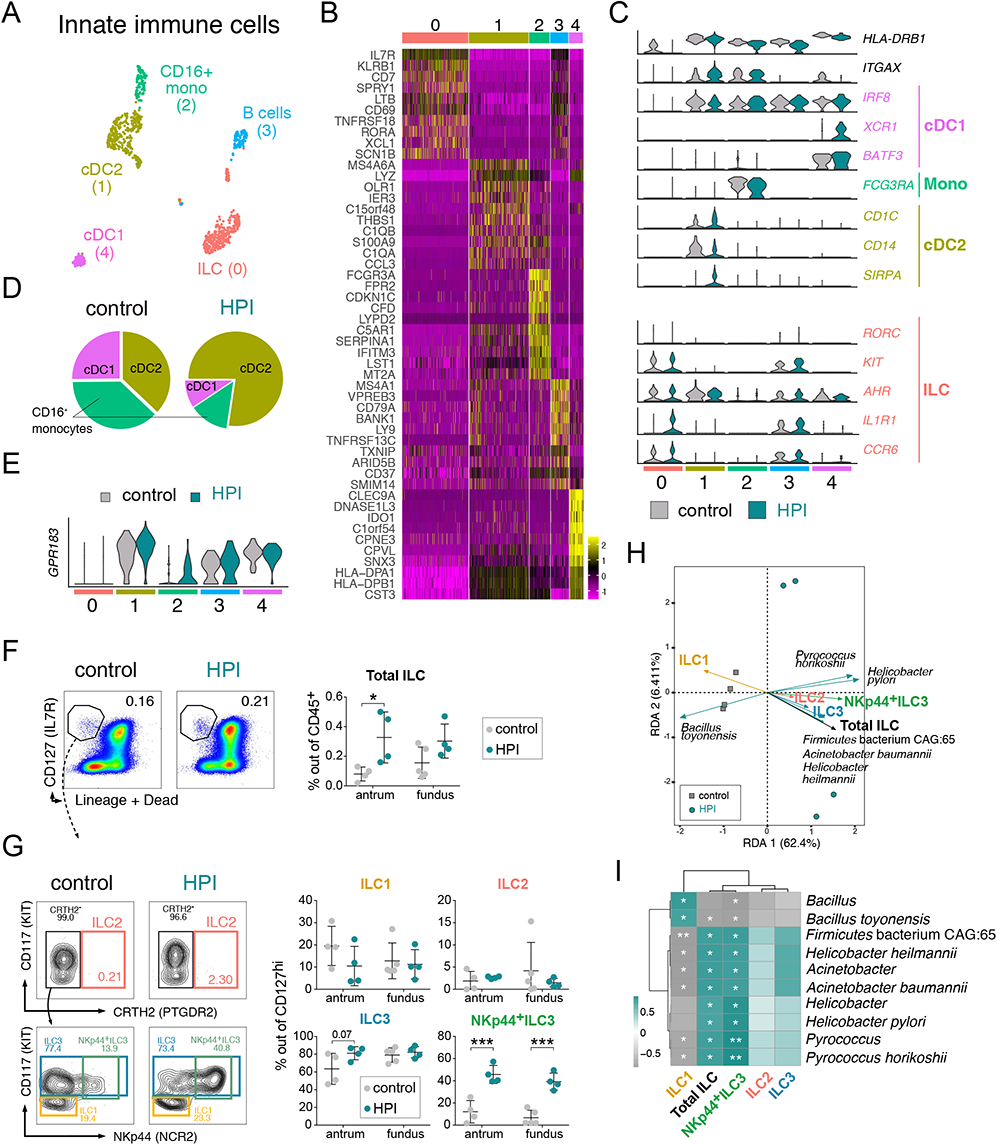
Innate immune cells from the HPI and uninfected tissues. **a.** UMAP showing unbiased clustering analysis of innate immune cells found in the gastric lamina propria. **b.** Heatmap of top differentially expressed genes between clusters shown in panel a. **c.** Violin plots showing expression of selected markers used for annotation of clusters in panel a. **d**. Frequency of each myeloid cell subset within total identified myeloid cells in the gastric lamina propria of HPI and uninfected tissues. **e**. Violin plots showing expression of *GPR183* gene. **f**. Representative flow cytometric plots (left) and quantification (right) of total ILC frequency out of CD45^+^ cells in the fundus and antrum of HPI and uninfected tissues, based on surface markers indicated in Table 3. **g.** Representative flow cytometric plots (left) and quantification (right) of ILC subset frequencies out of total ILC in the fundus and antrum of HPI and uninfected tissues, based on surface markers indicated in Table 3. **h.** Redundancy analysis (RDA) comparing ILC subset percentages from panel **g** with the relative abundance of microbial species that were differentially related to control or HPI tissues in figure 1e. **i**. Spearman correlation analysis between altered microbial species and ILC percentages induced by *H. pylori* infection. \**p* < 0.05, \*\**p* < 0.01.

### ILC3 are the dominant ILC population in the gastric mucosa

It has been reported that the ILC compartment of the murine stomach is predominantly composed of ILC2 (more than 90% of total ILC) with virtually no ILC3 ^11^. To further investigate this, we performed flow cytometric analysis of ILC subsets in the antrum and fundus of five uninfected and four HPI individuals. We found a significantly increased proportion of Lin^-^CD127^+^ ILC (out of total CD45^+^ cells) in the antrum of HPI as compared to uninfected individuals, with a similar trend, albeit not significant, in the fundus (figure 3F). As predicted by the gene expression profile, and in contrast to the mouse data ^11^, we observed low ILC2 and high ILC3 proportions in the human gastric mucosa, which were unaffected by *H. pylori* nor the region analyzed (figure 3G). *H. pylori* infection significantly increased the proportion of gastric NKp44^+^ ILC3 (figure 3G), which are known to play a role in epithelium defense and tissue-remodeling ^25^. To gain insights into the potential role of the gastric microbiome in affecting ILC homeostasis in the gastric mucosa, we performed redundancy analysis (RDA), which shows the correlation between the percentage of ILC measured by flow cytometry and the relative abundance of each differentially regulated microbial species from the same tissue. *H. pylori* and the bacterial species whose abundance increased upon infection (figure 1E) positively correlated with the proportion of ILC2, ILC3 and NKp44^+^ ILC3 (figure 3H, I). *B. toyonensis* showed an inverse correlation with ILC2 and ILC3, but a positive trend with ILC1 percentages (figure 3H, I). Correlation analysis indicates that the gastric microbiome and ILC responses are closely related and potentially influence each other. Altogether, in contrast to mice, ILC3 are the dominant ILC population in the human stomach and NKp44^+^ ILC3 are expanded in HPI individuals.

### Enhanced B cell activation in HPI gastric mucosa

We then focused on B cells and plasmablasts (*CD19*^+^*MS4A1*^+^, lacking *CD3D* and *ITGAX* expression) within the ILC & HLA-DR^+^ samples (figure 2D). Unsupervised clustering yielded 5 cell clusters annotated based on top differentially expressed genes (figure 4A-C). The predominant gastric B cell populations were *TNFRSF13B* (TACI)^+^ *GPR183*^+^ memory (cluster 0 and *YBX3*^+^ *CLEC2B*^+^ *TCL1A*^+^ naïve B cells (cluster 1). High expression of *HSP* family genes, *JUN* and *EGR1* identified cells in cluster 2 as activated B cells. Cells in cluster 3 were annotated as plasmablasts, based on high expression of *MZB1, XBP1,* and the endoplasmic reticulum (ER) chaperones *HSP90B1*, *DERL3* and *HSPA5* ^26^. Finally, we identified cells in cluster 4 as GC B cells, defined by their high expression of *RGS13*, *SUGCT*, *CD38* and *CXCR5* (figure 4B, C) ^27^. High expression of *AICDA* (AID) and *MSH6* ^28^, suggests that these cells are undergoing immunoglobulin (Ig) class switch recombination and somatic hypermutation (figure 4C). To further interrogate gene expression programs among gastric B cells in control and HPI mucosa, we applied topic modelling using latent Dirichlet allocations (LDA) ^29, 30^. Four out of six topics described major B cell gene expression programs within the gastric mucosa (figure 4D and suppl. figure 4A, B). Topic 1 predominantly weighted within cluster 0, and was defined by the expression of *TNFRSF13B*, *CLECL1* and *CRIP1* ^31, 32^ and high ribosomal genes, suggesting a relatively quiescent state, thus confirming the identity of memory B cells. Topic 2 heavily weighted within the cells annotated as naïve B cells and was defined, among other genes, by *YBX3*, *TCL1A* and *FCER2* expression. Topic 4 contained genes related to B cell activation, including *CD79B* and *JUN*. Although predominant in activated B cells (cluster 2), topic 4 was also shared by a fraction of naïve B cells (cluster 1) and memory B cells (cluster 0) (figure 3D), indicating the ability of both naïve and memory B cells within the gastric mucosa to undergo activation. Topic 6 was enriched in genes related to protein processing and immunoglobulin production (*PRDM1*, *CHST2*, *MZB1*, *FKBP11*) typical of plasmablasts ^33^, as well as genes associated with germinal center reactions (*BIK*, *MYBL1*) ^34, 35^.

**Figure 4.**
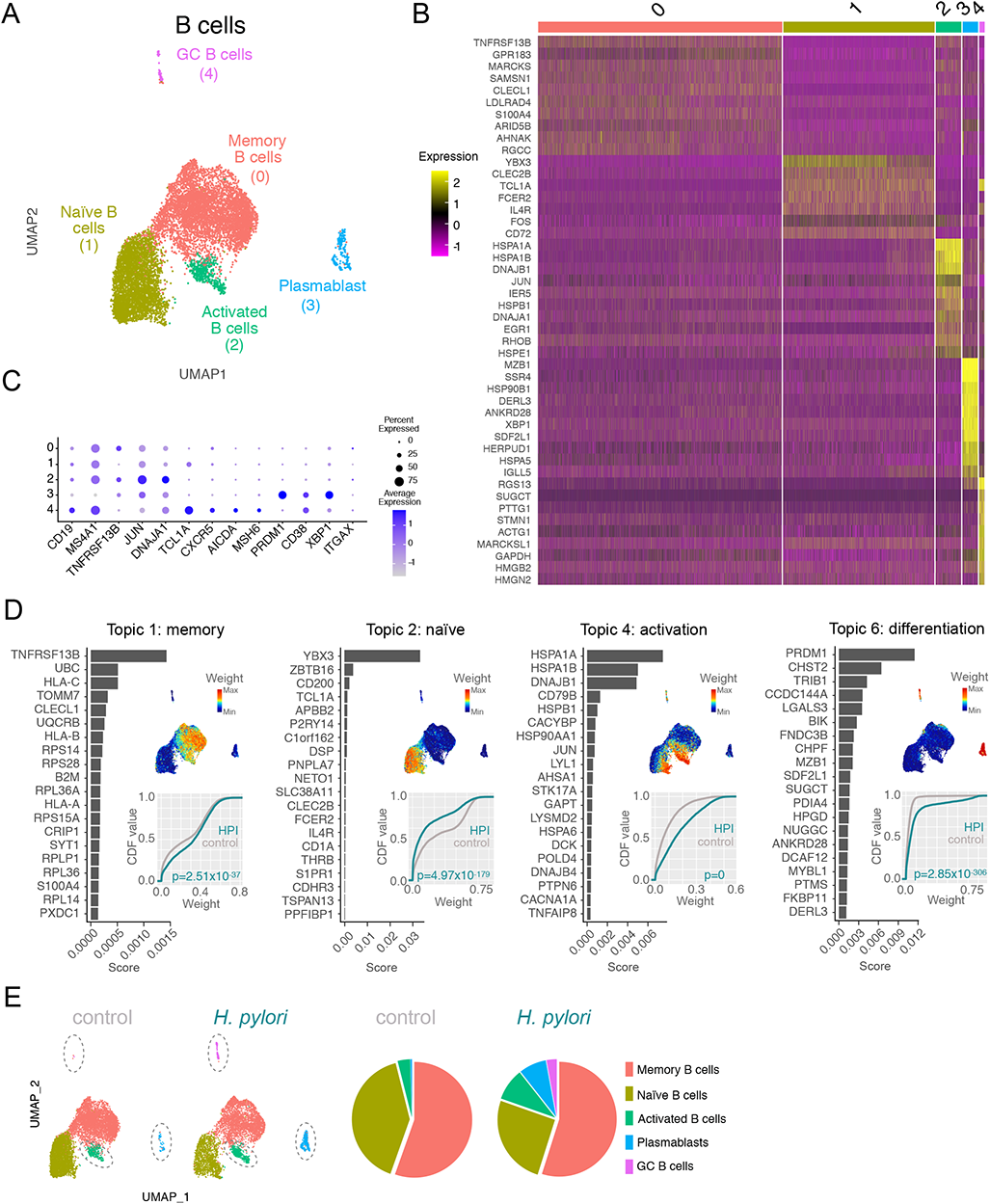
B cell activation and plasmablast differentiation are enhanced in HPI gastric mucosa. **a.** UMAP showing unbiased clustering analysis of B cell subsets found in the gastric lamina propria. **b.** Heatmap of top differentially expressed genes between clusters shown in panel a. **c.** Dotplot showing expression of selected cell markers that were used to annotate clusters in panel a. **d.** Topic modeling of B cell scRNA-seq data from HPI and uninfected tissues. This includes, for each topic: a bar plot showing the scores (x axis) of top-ranked genes within the indicated topic (left); a UMAP plot showing B cells colored by the topic’s weight in the cell (top right); the empirical cumulative density function (CDF, y axis) of topic weights (x axis) for the infected and uninfected condition (bottom right). Adjusted *p* values were determined using a Wilcoxon rank sum test. **e.** Frequency of each B cell cluster within total B cells in the gastric lamina propria of HPI and uninfected tissues.

To investigate differences in B cell activity between the HPI and uninfected gastric mucosa, we assessed the weight of these four topics in either condition. Topic 2 (naïve B cells) had a higher weight in cells from uninfected than HPI tissue. Conversely, topic 1, 4 and 6 (activated, differentiated and memory) were significantly more prominent in B cells from the HPI than uninfected gastric mucosa (figure 4D). Similarly, we detected an increased proportion of activated B cells, GC B cells and plasmablasts in the HPI as compared to uninfected sample (figure 4E, dashed ovals in Uniform Manifold Approximation and Projection, UMAPs). Together, these data indicate that *H. pylori* latent infection in asymptomatic individuals may promote the chronic activation of B cells, formation of germinal centers and differentiation of plasmablasts.

### *H. pylori* infection is associated with reduced CD8^+^ cytotoxic T lymphocytes (CTL) and increased T helper cells

We then analyzed the T cell compartment, sorted as Lin^+^ (suppl. figure 2A). Unsupervised clustering resulted in 6 distinct CD3^+^ T cell clusters (figure 5A-D and suppl. figure 5A). Clusters 0 and 3 were annotated as CTL based on the expression of *CD8A*, *CD8B*, *KLRD1* and *GZMB*. Additional expression of *GZMK* and *GZMA* suggested that cluster 3 may have an enhanced cytotoxic activity as compared to cluster 0 ^36, 37^. Cluster 5 grouped early activated CD4^+^ and CD8^+^ T cells characterized by high expression of *HSP*, *JUN*, *EGR1* and *CD69* (figure 5A-D). Clusters 1, 2 and 4 contained CD4^+^ T cells with different degrees of activation. Cluster 1 expressed *KLRB1*, *RORA*, *CCR6* and *IL22*, which pointed to pro-inflammatory T helper cells. Cluster 2 contained *CCR7*^+^ *SCML1*^+^ *SELL*^+^ naïve T helper cells. Cluster 4 comprised activated *CTLA4^+^* cells which expressed a mixed genetic signature pointing into either a Treg (*TNFRSF18*, *MAGEH1*, *TIGIT*) ^38, 39^ or follicular helper T (Tfh) direction (*CXCL13*, *BATF*, *IL6ST*) ^40–42^ (figure 5A-D). To assess functional differences between T cells in uninfected and HPI conditions, we applied topic modelling to this scRNA-seq dataset, focusing on topics whose weight differed significantly between the two conditions (figure 5E and suppl. figure 5B). Topic 1, characterizing a cytotoxic function with genes such as *KLRD1* and *GZMB*, was shared between the previously identified CTL clusters, and was more prominent in uninfected than HPI gastric mucosa. Topic 4 (naïve CD4^+^T cells) weight was also reduced in HPI samples. Conversely, topics 2 and 7, encompassing the CCR6^+^CD4^+^ T and CTLA4^+^CD4^+^ T clusters respectively, were more prominent in the infected condition (figure 5E). Having defined their gene expression profiles, we also compared proportions of cells within each cluster in control and HPI samples. We observed a decreased proportion of CTL1 and an increased proportion of CCR6^+^CD4^+^T and CTLA4^+^CD4^+^T cells in HPI compared to control individuals (figure 5E). Altogether, our data suggest an enhanced CD4 T helper response in the gastric mucosa of chronically HPI individuals compared to non-infected individuals.

**Figure 5.**
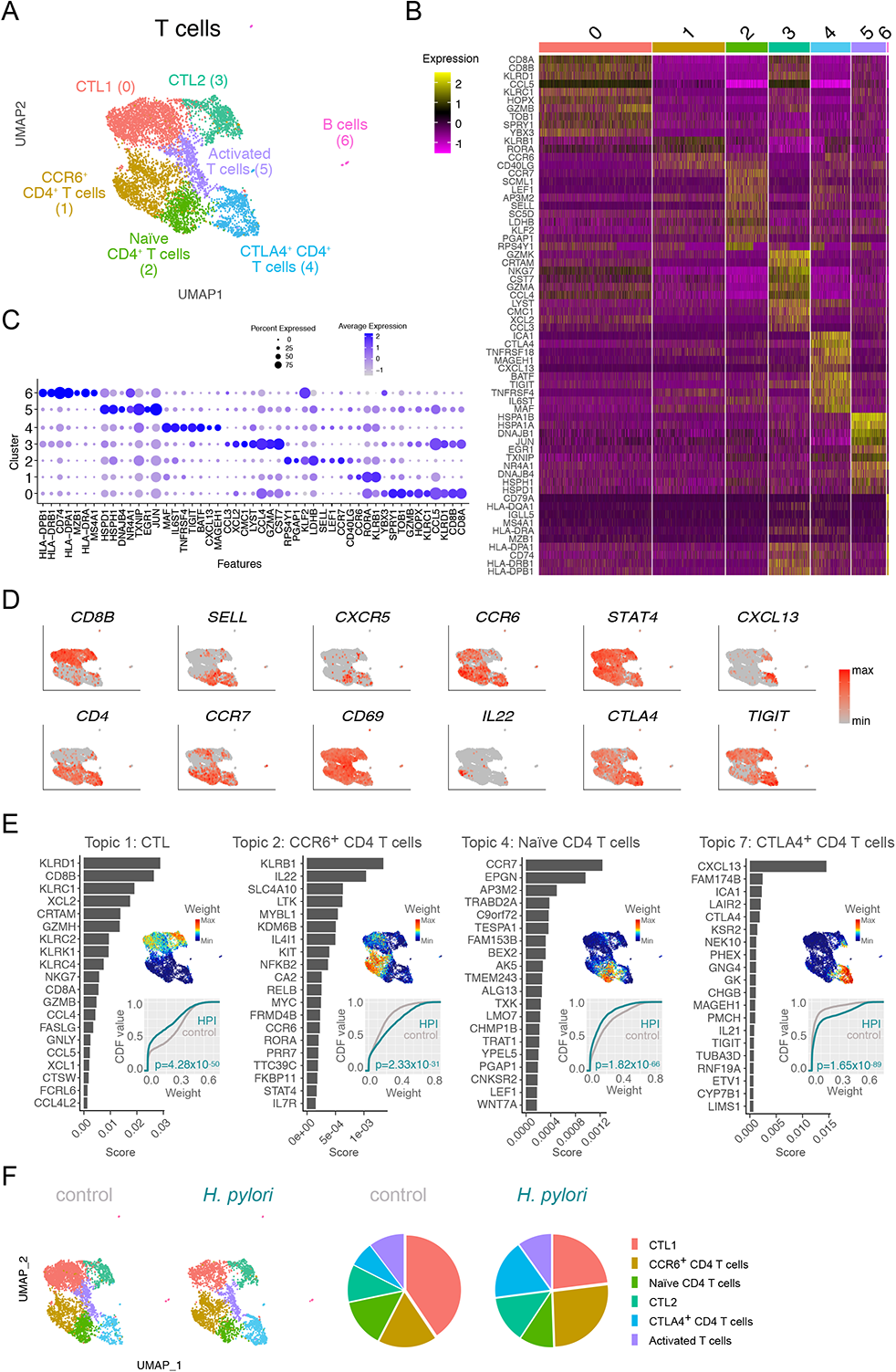
*H. pylori* infection is associated with reduced CD8^+^ cytotoxic T lymphocytes (CTL) and increased T helper cells. **a.** UMAP showing unbiased clustering analysis of T cell subsets found in the gastric lamina propria. **b.** Heatmap of top differentially expressed genes between clusters shown in panel a. **c.** Dotplot showing expression of selected cell markers that were used to annotate clusters in panel a. **d.** UMAP plots showing expression of selected genes used to annotate clusters in panel a. **e.** Topic modeling of T cell scRNA-seq data between HPI and uninfected tissues. This includes, for each topic: a bar plot showing the scores (x axis) of top-ranked genes within the indicated topic (left); a UMAP plot showing T cells colored by the topic’s weight in the cell (top right); the empirical cumulative density function (CDF, y axis) of topic weights (x axis) for the infected and uninfected condition (bottom right). Adjusted *p* values were determined using a Wilcoxon rank sum test. **f**. Relative abundance of each T cell subset within total T cells in the stomach of HPI and uninfected tissues.

### Chronic HPI individuals showed an enhanced Tfh program in the gastric mucosa

To further characterize the CTLA4^+^CD4^+^ T cell population, we subclustered it as shown in figure 6A, which resulted in five CD4^+^ T subclusters. Clusters 1 and 4 were characterized by high expression of Treg-related genes, including *FOXP3*, *IL10* and *LAG3* and they were annotated as IL10^+^Treg and Treg, respectively (figure 6A, B). Cluster 2 contained naïve T cells based on the expression of *KLF2* and *RTKN2* (suppl. figure 6A, B) ^43^. Clusters 0 and 3 were defined as Tfh cells based on the expression of *CXCL13*, *PDCD1*, and *CXCR5* (figure 6A, B). Analysis of subset proportion out of total T cells revealed that both Tfh and Treg cells increased upon *H. pylori* infection (figure 6C). Topic analysis further confirmed the follicular helper and regulatory activities of annotated T cells (figure 6D and suppl. figure 6C). A small portion of cells annotated as IL-10^+^ Treg cells, where *FOXP3* was not detected (figure 6B), co-expressed genes associated with a Th17-type immune response, including *IL17A*, *IL26* and *IL17F* (figure 6D, E) ^44^, which may hint at the activation of an IL-17 pathway as inferred by the microbiome data (figure 1F,G). Interestingly, the IL10^+^ Treg cluster also expressed high levels of *PTMS*, *ZNF282*, and *TP63* (figure 6E), which have been associated with oncogene activity and might help to explain the association of *H. pylori* infection with the development of gastric cancer ^45–47^. In summary, *H. pylori* infection correlates with a reduced CTL response in the gastric mucosa, accompanied by increased CD4^+^ T cell responses, which were characterized by gene expression programs associated with regulatory, oncogenic and Tfh cell functions.

**Figure 6.**
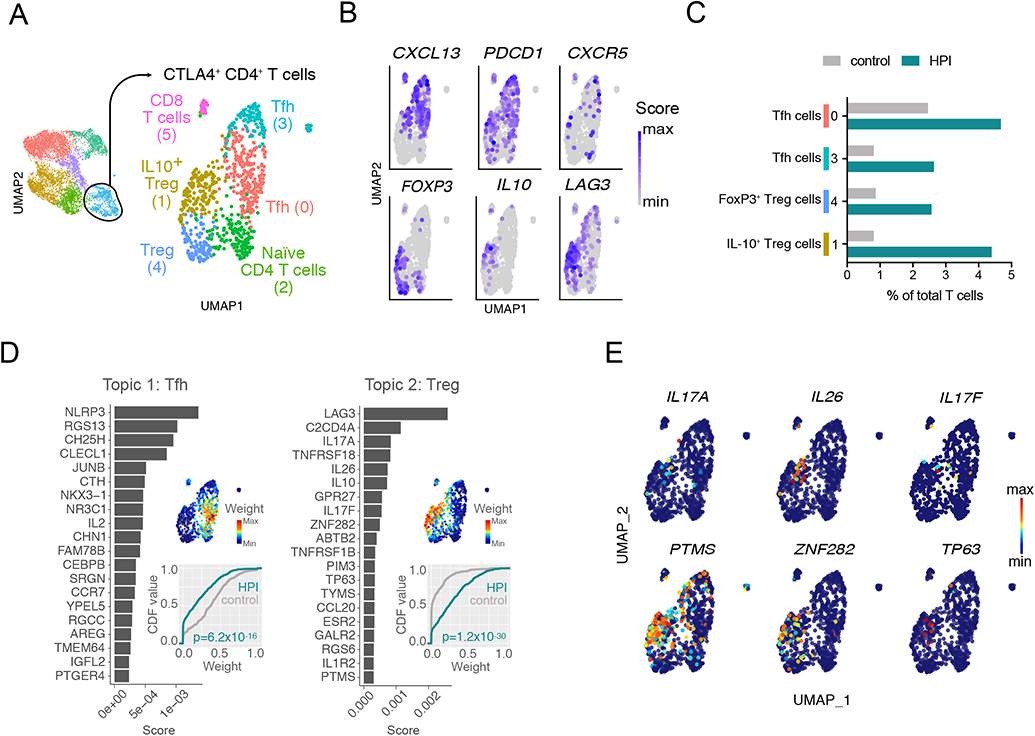
*H. pylori* infection is associated with increased regulatory T cells and T follicular helper cells. **a.** UMAP showing unbiased clustering analysis of CTLA4^+^CD4^+^ T cell subsets found in the gastric lamina propria. **b.** UMAP plots showing expression of selected genes used to annotate clusters in panel a. **c.** Frequency of subclusters of CTLA4^+^CD4^+^ T cells out of total T cells in HPI and uninfected tissues. **d.** Topic modeling of activated CD4^+^ T cell scRNA-seq data between HPI and uninfected tissues. This includes, for each topic: a bar plot showing the scores (x axis) of top-ranked genes within the indicated topic (left); a UMAP plot showing T cells colored by the topic’s weight in the cell (top right); the empirical cumulative density function (CDF, y axis) of topic weights (x axis) for the infected and uninfected condition (bottom right). Adjusted *p* values were determined using a Wilcoxon rank sum test. **e.** Weight of selected genes that define topic 2 in panel d.

### Innate and adaptive immune cells interact to form gastric tertiary lymphoid structures (TLS) during asymptomatic H. pylori infection

Gastric ILC, cDC2 and B cells showed high *LTB* and *GPR183* levels (figure 3B, E and figure 4B), which pointed to the formation of mucosal TLS ^48, 49^. Human mucosa-associated TLS are mainly composed by activated/GC B and Tfh cells ^50^, whose relative abundance was increased in the HPI gastric mucosa. Therefore, we hypothesized the existence of mature TLS in HPI individuals. To test this, we first inferred paracrine interactions between the identified cell types using SingleCellSignalR analysis of transcriptomic data ^51^. We quantified the putative ligand-receptor (LR) pairs between B cells, T cells and innate cell types. While the overall number of putative interactions with LR score>0.5 was similar between control and HPI gastric mucosa, 63 interactions (17% of total) only occurred upon *H. pylori* infection and 75 interactions (21%) were specific to the uninfected gastric mucosa (figure 7A). Gene Ontology (GO) enrichment analysis of LR pairs indicated that pathways associated with the uninfected mucosa were mostly related to innate immune function and T cell tolerance. Conversely, *H. pylori* infection enhanced pathways related to cytokine production and leukocyte aggregation (figure 7B). Most putative interactions observed in the HPI stomach tissue involved myeloid cells as either ligands or receptors. GC B cells and CTLA4^+^CD4^+^ T cells offered the highest number of receptors in the B cell and T cell fractions respectively (figure 7C). In addition, ILCs, cDC2 and CTLA4^+^CD4^+^ T cells from the HPI (but not control) gastric mucosa were predicted to interact via the *LTB*-*LTBR* axis and *VEGFA* angiogenic signals (suppl. figure 7A,B) ^52^. Therefore, analysis of putative paracrine interactions predicts the possibility that the identified cell types interact with each other to form TLS in the HPI gastric mucosa. To validate the interaction of B cells, T cells and CD11c^+^ myeloid cells within TLS in the gastric mucosa, we performed an immunofluorescent co-staining of CD20, CD3 and CD11c on uninfected and HPI tissues. All HPI samples analyzed (n=3 distinct individuals), but not control samples, showed leukocyte aggregates, usually characterized by a CD20^+^ B cell area surrounded by a CD3^+^ cell region and interspersed CD11c^+^ cells (figure 7D), indicating the formation of TLS in asymptomatic HPI individuals. Thus, transcriptomic and immunofluorescent data point towards the formation of mature TLS as a hallmark of chronic *H. pylori* infection.

**Figure 7.**
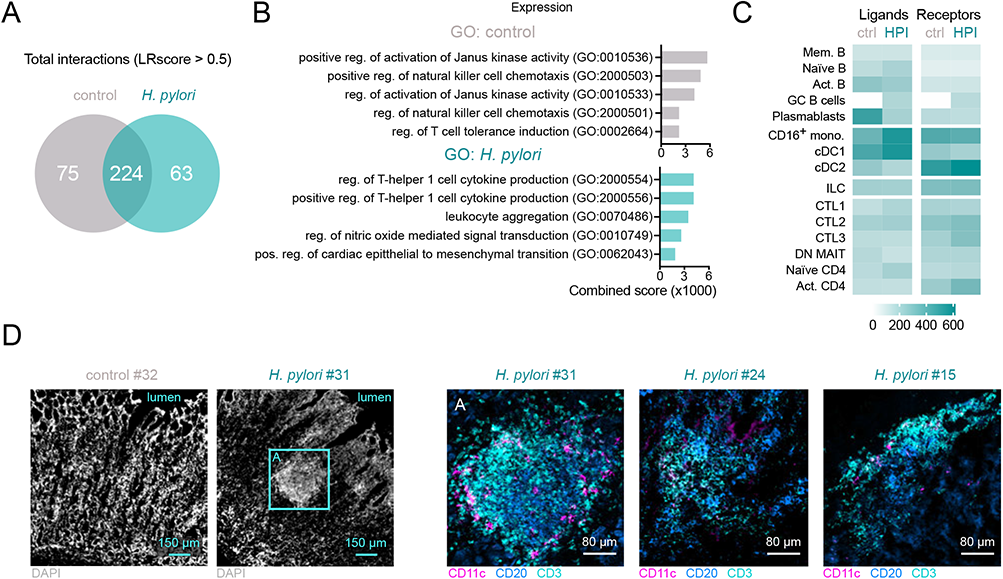
Innate and adaptive immune cells interact to form gastric tertiary lymphoid structures (TLS) during asymptomatic H. pylori infection. **a.** Number of ligand-receptor interactions with score > 0.5 predicted by SingleCellSignalR analysis of transcriptomic data of all cell subsets identified in Figure 3a, 4a and 5a, in HPI or uninfected tissues. **b.** Gene ontology (GO) enrichment analysis of ligands and receptors involved in predicted interactions in panel a. **c.** Number of putative ligands and receptors offered by any cell type involved in interactions in HPI or uninfected tissues. **g.** Multicolor immunofluorescence microscopy demonstrating distribution of CD3^+^, CD20^+^ and CD11c^+^ cells within the gastric mucosa of HPI and uninfected tissues.

## DISCUSSION

By combining microbiome sequencing, scRNA-seq, flow cytometry and immunofluorescent microscopy from human gastric tissues, our findings expand current knowledge of the gastric microbiome and immune cell function in uninfected and HPI mucosa. We showed that asymptomatic *H. pylori* infection severely alters the microbiome composition and functional profile in the stomach mucosa. Using single-cell RNA sequencing (scRNA-seq) and flow cytometry, we provided a high-resolution characterization of the human gastric immune cell composition.

As nearly all previous studies on the gastric microbiome were obtained via 16S rRNA gene sequencing, our use of whole genome sequencing provides more information at the species level and a glimpse of the corresponding bacterial function. *E. coli* and *S. pneumoniae,* the predominant species comprising the gastric microbiome in control samples, are often found in other studies ^5, 6^. Also in line with previous studies, we demonstrated a significant shift of the gastric microbiome composition after *H. pylori* infection ^6, 53, 54^. Moreover, we observed a significant increase of the genes encoding for enzymes involved in LPS and glycan biosynthesis and metabolism within the HPI tissues. LPS has been shown to be actively involved in the immune response via modulation of antigen processing and presentation ^55^, the Th17 pathway ^56^, and the NOD-like receptor signaling pathway ^57^. Glycans also serve as key checkpoints of T cell function ^58^. These findings are further supported by our metagenomic pathway analysis and corresponding immune survey. Although the high predominance of *H. pylori* may suggest it is the lead player in eliciting immune activation and subsequent gastritis, the changes in the immune response may also result from the alteration of the whole gastric microbiome. The correlation between gastric microbes other than *H. pylori* and immune pathways provided evidence for this hypothesis and further prompted us to investigate the effect of such a dysbiotic microenvironment on the immune compartment in the underlying gastric mucosa.

Our scRNA-seq analysis of the B cell compartment revealed that *H. pylori* infection promoted B cell (re)activation, GC formation and differentiation of plasmablasts, which were hardly detectable in uninfected tissues. This is in line with previous reports of B cell follicle formation and plasma cell accumulation in the gastric lamina propria of HPI patients and *Helicobacter* spp.-infected mice ^59, 60^.

We described a shift in the balance between CD8^+^ CTL and CD4^+^ T cells in HPI tissues as compared to uninfected tissues. An enhanced Tfh cell function is in line with the increased GC B cells detected in the infected tissues. Notably, activation of the Tfh program is not impaired, but rather coexists with the immunosuppressive function of Treg cells, that have previously been linked to evasion of *H. pylori* from immune-mediated clearance mechanisms and protection from peptic ulcer ^61–63^. Our observations thus support the hypothesis of an equilibrium between immunosuppressive environment and B cell expansion in the establishment of asymptomatic *H. pylori* infection in the human gastric mucosa, as was previously suggested in mice ^64^. While decreased CTL function fits an immunosuppressive environment, understanding the potential role of increased CCR6^+^CD4^+^ T cells upon *H. pylori* infection requires additional insight.

Numerous studies have investigated the immune response against persistent microbial stimulation of the intestinal mucosa and the formation of TLS in the intestine, but rarely in the human stomach. Interestingly, the transcriptional profile of cDC2 that were predominant among myeloid cells in the HPI gastric mucosa resembled the one of DCs found in association with intestinal cryptopatches ^65^. Similarly, ILC3 have been previously shown to promote mucosal antibody responses in the murine intestine ^66, 67^, and we also found that NKp44^+^ ILC3 were increased in HPI as compared to uninfected individuals. These cells are known to play an important role in epithelium defense and tissue-remodeling in the small intestine ^25^, and further studies are needed to assess whether they could exert a similar function in the gastric mucosa.

This is to our knowledge the first characterization of the ILC compartment in the human gastric mucosa by both scRNA-seq and flow cytometry. An elegant study has recently reported that the murine stomach contains predominantly ILC2 cells, which promote *H. pylori*-specific IgA responses upon infection ^11^. The apparent discrepancy between this murine study and our observations in human tissue could be explained by several factors, including acute vs chronic type of the infection, host variation, microbiome and diet differences, and other environmental exposures. Moreover, ILC2 have been recently shown to undergo transition to an ILC3-like state under certain inflammatory conditions, which would support a less compartmentalized description of the ILC of the stomach ^30, 68^.

One limitation of the current study is low sample size. Despite this, we performed multiple investigative approaches on the same human gastric tissues, which provide novel correlations and predict interactions between gastric microbiome and immune cells. Secondly, the tissues were obtained from obese individuals, which might influence the findings of gastric microbiome and immune response. However, our data were generated by comparing tissues with and without *H. pylori*, which should minimize the confounder factor derived from obesity. In addition, this is not a complete immune cells survey, as we left out some immune cells, such as granulocytes and NK cells, from scRNA-seq analysis. Further studies are needed to prove if *H. pylori* infection is the cause for all the changes observed in the gastric microbiome and immune cells.

## METHODS

### Study population

In total 24 samples were obtained from 14 individuals undergoing sleeve gastrectomy for morbid obesity at Ersta hospital in Stockholm, Sweden. The samples were kept on ice and processing started within 40 min from the surgery. The *H. pylori* infection status of each stomach sample was detected using BIOHIT *H. pylori* quick test kit (Biohit HealthCare, United Kingdom) on two representative pieces of stomach tissue from fundus and antrum. *H. pylori* status was further determined by culturing swab samples from 28-32 sites of the stomach tissues on GC agar plates (Karolinska Hospital, MIK0346, Sweden) under microaerophilic condition. Four individuals were defined as the *H. pylori*-infected and eight were *H. pylori*-uninfected. Detailed characteristics of the participants, based on questionnaire data, are listed in Table 1. The study was approved by the Regional Ethical Board at Karolinska Institute, Stockholm, Sweden.

**Table 1.** Clinical characteristics of all patients.

| Patient | Age range | Height | Weight | Gender | *H. pylori* infection | Experimental purpose |  |  |  |
| --- | --- | --- | --- | --- | --- | --- | --- | --- | --- |
|  |  |  |  |  |  | FACS | Immuno-fluorescence | Microbiome seq ID | scRNA seq. |
| 01 | 51-60 | 167 | 121 | Woman | Positive | X |  |  | X |
| 02 | 21-30 | 172.5 | 98 | Woman | Negative | X |  |  | X |
| 03 | 21-30 | 176 | 130 | Woman | Negative | X |  |  |  |
| 04 | 31-40 | 161 | 94 | Woman | Positive | X | X |  | X |
| 05 | 61-70 | 162 | 90 | Woman | Negative | X |  |  |  |
| 06 | 31-40 | 161 | 103 | Woman | Negative |  | X |  |  |
| 07 | 31-40 | 189 | 150 | Man | Negative | X |  | Control 1 | X |
| 08 | 31-40 | 160 | 89 | Woman | Positive | X | X | *H.pylori* 1 |  |
| 09 | 31-40 | 171 | 114 | Woman | Negative | X | X | Control 2 | X |
| 10 | 31-40 | 165 | 104 | Woman | Negative |  |  |  |  |
| 11 | 61-70 | 150 | 87 | Woman | Positive | X | X | *H.pylori* 2 | X |
| 12 | 41-50 | 173 | 115 | Woman | Negative |  | X | Control 3 | X |
| 13 | 21-30 | 164 | 102 | Woman | Negative |  |  |  | X |
| 14 | 21-30 | 165 | 129 | Woman | Negative |  |  |  | X |

Epithelial microbiome samples were collected from fundus and antrum using swabs (FLOQSwabs, Copan Flock Technologies, Italy), put directly into DNA/RNA shield (Zymo Research, USA) and snap-frozen at −80°C until further DNA extraction and metagenome sequencing. Two pieces of gastric lamina propria tissues from fundus and antrum were processed for isolation of mononuclear cells. In addition, tissues (around 1 × 1 cm^2^) from fundus and antrum were collected and snap-frozen for multicolor immunofluorescence microscopy.

### Metagenome sequencing

Stomach microbiome samples (n=10, four *H. pylori*-infected and 6 *H. pylori* uninfected by *H. pylori* quick test kit and culture) were bead-beaten with Matrix E beads (MP Biomedicals, USA) for three times of two minutes. The samples were then incubated with 20 µl of lysozyme (100 mg/ml, Sigma-Aldrich) at 37°C for 60 min followed by digestion with 20 µl proteinase K (20 mg/ml, Qiagen) at 55°C, 250rpm for 90 min. The following steps were performed with DNeasy blood & tissue kit (Qiagen, USA) according to the manufacturer’s guidelines. The following metagenomic sequencing and analysis was carried out at Novogene located in the UK. Briefly, extracted DNA was fragmented to an average size of about 400 bp using Covaris M220 (Gene Company Limited, China) for paired-end library construction using NEXTFLEX Rapid DNA-Seq (Bioo Scientific, Austin, TX, USA). Adapters containing the full complement of sequencing primer hybridization sites were ligated to the blunt end of fragments. Sequencing was performed on Illumina NovaSeq/Hiseq Xten (Illumina Inc., San Diego, CA, USA) using NovaSeq Reagent Kits/HiSeq X Reagent Kits according to the manufacturer’s instructions.

### Analysis of Metagenome sequencing data

Reads that mapped to the human genome together with their mated/paired reads were removed from each sample using soap 2.21 ^69, 70^. The quality control of each dataset was performed to trim low quality bases (Q-value ≤ 38) which exceed 40 bp, reads containing ambiguous nucleotide base calls (“N”) in tandem sequences ≥ 10 bp, and reads overlapping with adapter sequences at lengths over 15 bp. Samples passing quality control (QC) were assembled initially using an optimized SOAPdenovo protocol to obtain the scaftigs without “N” ^71, 72^. Clean data were mapped to assembled Scaftigs using Soap 2.21 and unutilized Paired-end reads were collected ^73^. Mixed assembly was conducted on the unutilized reads with the same assemble parameter and the mixed assembled scaftigs below 500 bp were trimmed ^70, 72, 74^. The effective Scaftigs were used for further analysis and gene prediction. MetaGene (http://metagene.cb.k.u-tokyo.ac.jp/) was used to predict open reading frames (ORFs) from the assembled contigs with the length over 100 bp ^75^. Assembled contigs were then pooled and non-redundancies were constructed based on the identical contigs using CD-HIT with 95% identity (http://www.bioinformatics.org/cd-hit/) ^76^.

Original sequences were mapped to the predicted genes and estimate the abundances using SOAPaligner (http://soap.genomics.org.cn/) ^77^. Taxonomic assessment of stomach microbiota was performed using MEGAN ^78^ against the sequences of Bacteria, Fungi, Archaea, and Viruses extracted from NR database (NCBI: version 2016-11-05) (http://www.ncbi.nlm.nih.gov/RefSeq/). Taxonomic profiles were conducted at the domain, phylum, genus and species levels, with relative calculated abundance. Very low abundance of *Candidatus.Kryptonium.thompsoni*, which was found at the same level in all the samples, was considered as contamination and removed from the analysis. The principal coordinates analysis (PCoA) based on Bray-Curtis dissimilarity matrices at genera and species level was performed. Contigs were annotated using DIAMOND against the Kyoto Encyclopedia of Genes and Genomes (KEGG) database (http://www.genome.jp/kegg/) with an E value of 1e-5 [doi: 10.1093/nar/28.1.27.]. The Carbohydrate-Active enZYmes (CAZy) Database annotation was performed using USEARCH (http://www.drive5.com/usearch/), and antibiotic resistance gene analysis was performed by querying gene sequences against the CARD (the Comprehensive Antibiotic Research Database) database ^79, 80^. Abundances of KEGG Orthology (KO), pathway, KEGG enzyme, Module, and CAZymes were normalized into counts per million reads (cpm) for downstream analysis. The differential taxonomic and metagenomic function results were obtained using Metastats analysis with q value < 0.05 ^81^ or LEfSe (linear discriminant analysis (LDA) Effect Size) analysis with p value < 0.05 and LDA scores> 4 ^82^.

### Isolation of mononuclear cells from the gastric lamina propria

Gastric tissue (c.a. 1-5 g) was cleaned from fat, cut into 0.5×1 cm pieces. Tissues were incubated for 30 min at 37 °C in 20 ml Hank’s balanced salt solution (HBSS) with 5% fetal calf serum (FCS), 5 mM ethylenediaminetetraacetic acid (EDTA), 1 mM dithiothreitol (DTT) and 15 mM 4-(2-hydroxyethyl)-1-piperazineethanesulfonic acid (HEPES) under gentle shaking. The tissues were washed in 20 mL PBS with 5% FCS and 1 mM EDTA at 37 °C followed by washing in PBS with 1% FCS and 15 mM HEPES at room temperature. Tissues were then digested in 10 ml of serum-free HBSS with Collagenase D (0.5 mg/ml, Roche) and 0.1 mg/ml DNase I (Roche) at 37 °C, 600 rpm for 35 min followed by filtration through a 100 μm cell strainer. A 44–67% Percoll (Sigma Aldrich) separation was performed to enrich mononuclear cells. Cells were immediately used or frozen in FCS + 10% dimethyl sulfoxide (DMSO) for later use.

### Cell sorting prior to scRNA library preparation

Flow cytometry-activated cell sorting (FACS) was used to select gastric immune cell populations. Frozen cells that had been previously isolated from the lamina propria of the fundus and antrum were thawed in a 37°C water bath. Samples from the same group (region of the stomach, *H. pylori* presence) were pooled and washed in warm complete RPMI-10%FCS. After washing, cells were counted and resuspended in PBS supplemented with 2 mM EDTA and 5% FCS (sorting buffer). Cells were then stained with the appropriate hashtag antibody (B1 for fundus and B2 for antrum) and identical staining antibodies (Table 2) for 30 min at 4 °C, before being washed 3 times in sorting medium. Cells were then resuspended in sorting buffer with 4′,6-diamidino-2-phenylindole (DAPI) and sorted on a SH800S sorter (Sony) with “Purity” sorting mask. Cells were sorted as single DAPI^-^ CD45^+^Lineage^+^ (T cells), DAPI^-^CD45^+^Lineage^-^CD127^+^ (ILC) and DAPI^-^CD45^+^Lineage^-^ CD127^-^HLA-DR^+^ (B cells and myeloid cells). After cell counting, cells were pooled as specified below for single-cell RNA isolation.

**Table 2.**
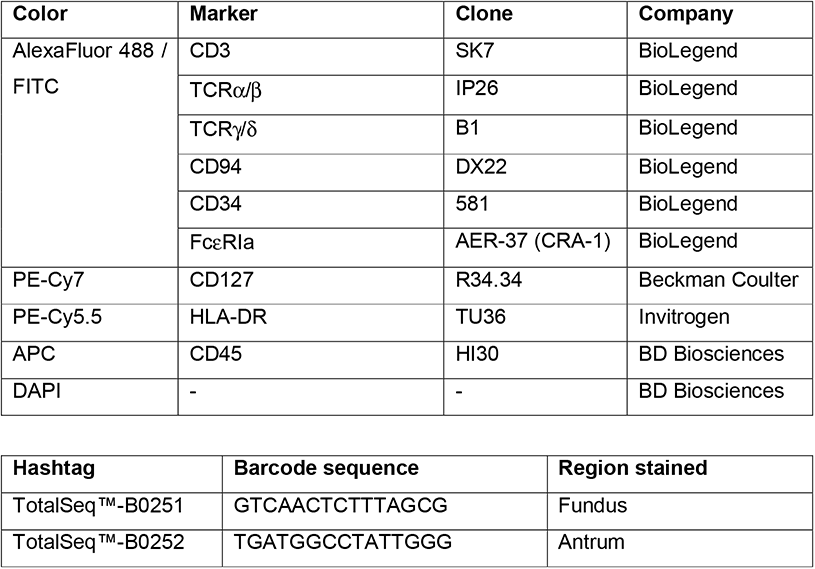
Antibodies used for cell sorting prior to scRNA isolation.

### Single-cell RNA isolation and sequencing

After Trypan blue counting, equal amounts of T cells from the fundus and antrum were pooled according to their *H. pylori* status (positive or negative). Similarly, equal amounts of B cells & myeloid cells from the fundus and antrum samples were pooled according to their *H. pylori* status (positive or negative). All ILC from the antrum and fundus were pooled according to their *H. pylori* status (positive or negative). T cells were loaded on two wells of a chip B (10X genomics) according to manufacturer instructions; a mix of myeloid, B cells and ILCs were loaded on two other wells of the same chip, for a total of four wells. Samples were processed with Chromium Single Cell 3**’** Reagent Kits v3 with Feature Barcoding technology for Cell Surface Protein (10X genomics) according to manufacturer instructions. Samples were sequenced at Novogene, UK.

### Analysis of single-cell transcriptomic sequencing (scRNA-seq) data

To analyze the single cell data obtained from fundus and antrum region of the stomach, we employed 10X genomics Cell Ranger 3.1.0 pipeline for preprocessing the data, Seurat 3.1.3 R package ^83^ for downstream single cell clustering and differential expression analysis. Single cell SignalR package ^51^ for potential cell-cell interaction analysis.

First, we aligned the reads from stomach single cell datasets on the reference hg19 and Grch38 (hg38) genomes using cell ranger pipeline, for the quantification of the cells as well as genes and cell surface proteins. We follow the quality check and filtering protocol based on cutoff criteria for cells (no. of feature expression per cell > 200 and < 2500); gene expression (>= 3 cells), mitochondria content (< 10 %), ribosomal content (> 5%), regression of cell cycle effect and removing the non-coding gene from the expression dataset. After filtering out cells through proper quality control procedure, we ended up with 22033 cells with 19774 genes expressed in total. For downstream analyses including dimensionality reduction, clustering and markers identification, we used the Seurat 3.1.3 package in our analysis pipeline. We first clustered all the cells together based on gene expression as well as cell surface antibody and obtained the Uniform Manifold Approximation and Projection (UMAP) and t-distributed stochastic neighbor embedding (t-SNE) embeddings. We generated the heatmap and dot-plot based on expressed marker genes (*p* <= 0.05, min.pct = 0.25, logfc.threshold=0.25) in each cluster. Based on canonical markers expressed in each cluster, we further subset the single cell dataset into B cells, ILC/myeloid and T cells and reanalyzed them separately to obtain further cell specific subclusters. This allowed us to determine differential expression of genes (*p* <= 0.05, min.pct = 0.25, logfc.threshold=0.25) as well as cell specific sub-population differences between control and the HPI condition. As an alternative approach to determine the difference between the control and HPI condition, we followed a topic modeling protocol ^29^. Using this approach, we obtained multiple topics (grade of membership models, in which individual cells represent partial membership in several sub-populations of cells simultaneously, based on the shared transcriptional rewiring) and their corresponding weightage difference between control and HPI condition. Further on, to understand the cell-cell interactions between different types of cells in the stomach single cell dataset, we ran SingleCellSignalR analysis to obtain ligand-receptor interactions between different types of subcellular population in the control vs HPI condition.

### Multicolor immunofluorescence microscopy

Human stomach biopsies were embedded in Tissue Tek O.C.T. medium (Sakura Finetek) and snap frozen with dry ice before storage at −80°C. 8 µm sections were cut using a cryostat and mounted on Superfrost plus glass slides (ThermoFisher Scientific). All slides were kept at −80°C. Staining was performed as described elsewhere ^84^, with minor modifications. Prior to staining, the tissue sections were air-dried for 15 min and fixed with 2% PFA in PBS for 20 min. The sections were permeabilized and blocked in Tris-buffered saline with 4% FCS, 4% BSA, 1% HEPES and 0.1% saponin for 30 min. The avidin/biotin blocking kit (Vector Laboratories) was used to block endogenous biotin. Sections were stained over night at 4°C with polyclonal rabbit anti-human CD3 (Dako), biotinylated monoclonal mouse anti-human CD20 (BD Biosciences, clone 2H7), and PE-conjugated monoclonal mouse anti-human CD11c (BD Bioscienes, clone B-ly6). Sections were subsequently blocked with 1% donkey serum and incubated with A488-conjugated donkey anti-rabbit (Invitrogen) and streptavidin-A647 (Biolegend) for 45 min at RT. All slides were mounted with Fluoromount Aqueous Mounting Medium (Sigma-Aldrich) and images were acquired on an LSM 700 system (Carl Zeiss) equipped with 405-, 488-, 555-, and 639 nm excitation lines at the Biomedicum Imaging Core at the Karolinska Institute. Image analysis was performed using Zen 2.3 software (Carl Zeiss) or Imaris Viewer 9.5.1 (Bitplane).

### Flow cytometry

For flow cytometry analysis of ILCs, frozen isolated mononuclear cells were thawed, washed extensively in PBS supplemented with 2 mM EDTA and 5% FCS and stained with antibodies (Table 3) for 20 min in at 4 °C. Cells were washed and acquired on an LSR Fortessa flow cytometer (BD Biosciences). Data were analyzed with FlowJo software (Tree Star) and statistical differences between multiple groups were assessed by 2-way ANOVA.

**Table 3.** Antibodies used for immunophenotyping of ILCs.

| **Color** | **Marker** | **Clone** | **Company** |
| --- | --- | --- | --- |
| AlexaFluor 488 / FITC | CD3 | SK7 | BioLegend |
|  | CD19 | 4G7 | BD Biosciences |
|  | CD14 | TÜK4 | Dako |
|  | CD1a | HI149 | BioLegend |
|  | CD123 | 6H6 | BioLegend |
|  | BDCa2 | AC144 | Miltenyi |
| | TCR$\alpha/\beta$ | IP26 | BioLegend |
| | TCR$\gamma/\delta$ | B1 | BioLegend |
|  | CD94 | DX22 | BioLegend |
|  | CD34 | 581 | BioLegend |
| | FcεR1a | AER-37 (CRA-1) | BioLegend |
|  | Live/Dead | - | Invitrogen |
| PE-Cy7 | CD127 | R34.34 | Beckman Coulter |
| PE-Cy5.5 | CD117 | 104D2D1 | Beckman Coulter |
| PE-Cy5 | Nkp44 (CD336) | Z231 | Beckman Coulter |
| PE-CF594 | CRTH2 (CD294) | BM16 | BD Biosciences |
| APC | NKG2a (CD159a) | Z199 | Beckman Coulter |
| QD605 | CD161 | HP-3G10 | BioLegend |
| AmCyan | CD45 | HI30 | BD Biosciences |

### Data Availability Statement

Nucleotide sequencing data are made available in public, open access repositories. Shotgun metagenomic DNA sequences were deposited to the European Nucleotide Archive (ENA) under project accession number PRJEB48730. scRNA-seq data were submitted to the European Nucleotide Archive (ENA) under project accession number PRJEB48798.

## ABBREVIATIONS

GC: germinal center
HPI: *H. pylori*-infected
Tfh: follicular helper T cell
Treg: regulatory T cell

## ACKNOWLEDGEMENTS

J.D. was supported by the Swedish Foundation for Strategic Research (SSF) (ICA16-0050), Svenska Läkaresällskapet (SLS-960584) and Karolinska Institutet. A.T. was supported by Erling-Persson Foundation (140604) and L. E. was supported by Söderbergs foundation. E.J.V. was supported by grants from the Swedish Research Council, VR grant (K2015-68X-22765-01-6 and 2018-02533), Formas grant (FR-2016/0005), Cancerfonden (19 0395 Pj), and the Wallenberg Academy Fellow program (2019.0315). The computations and data handling were enabled by resources provided by the Swedish National Infrastructure for Computing (SNIC) at KTH partially funded by the Swedish Research Council through grant agreement no. 2018-05973. Ferring Phamaceuticals funded the centre where the sample collection and microbiome related experiments were carried out.

---

**Suppl. Figure 1.**
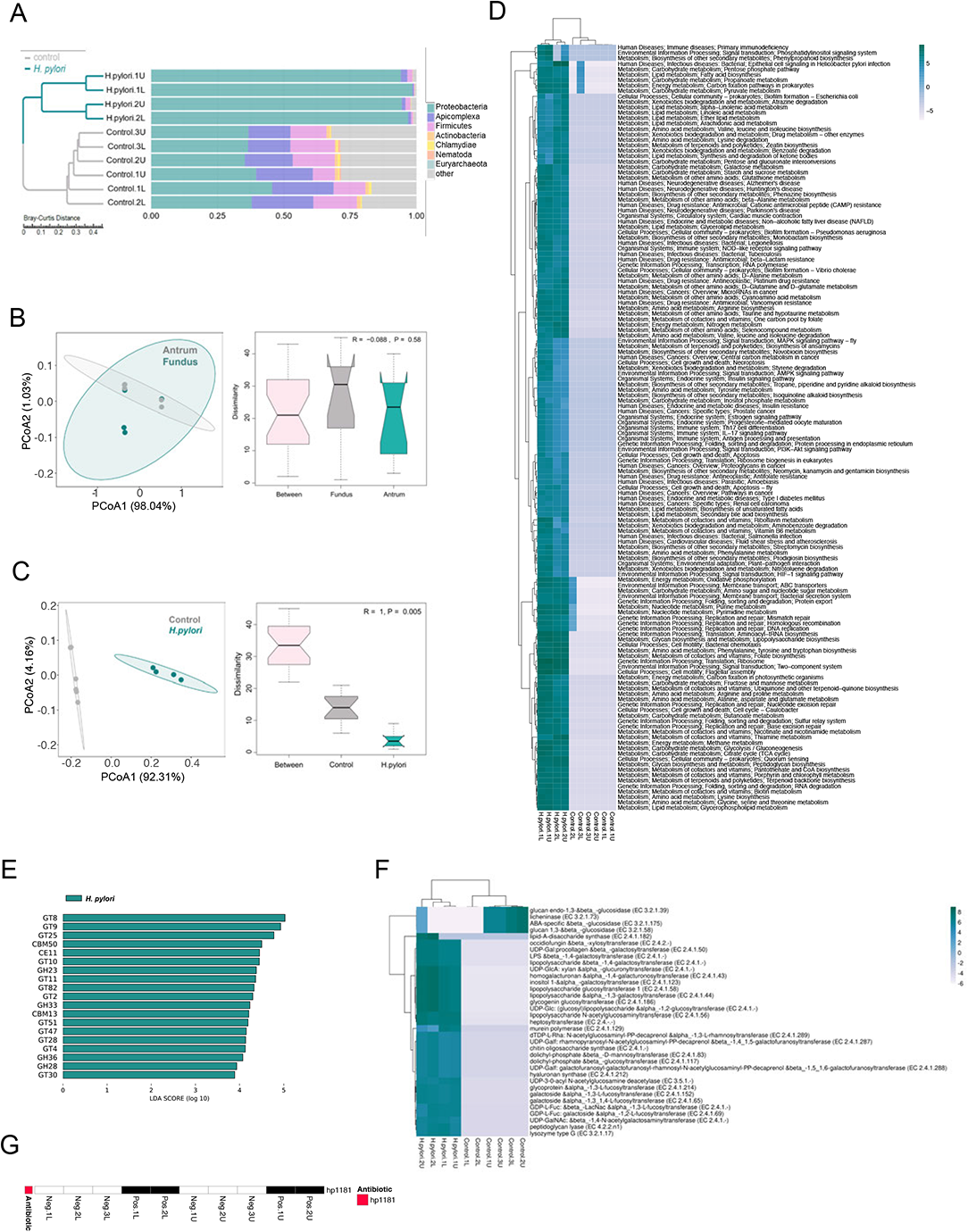
Composition and function of the gastric microbiome are altered in H. pylori-infected tissues. **a.** The ten most abundant bacteria at the Phylum level and sample clustering of the HPI and uninfected tissues based on Bray-Curtis distance. **b.** The PCoA plot of fundus and antrum samples with identified bacterial species based on Bray-Curtis distance. The analysis of similarities (Anosim) showing no difference in the microbial diversity between the fundus and antrum regions. **c.** The PCoA plot based on functional abundance in KEGG database. The analysis of similarities (Anosim) showed a significant difference in the functional abundance of the HPI and uninfected tissues. **d.** Heatmap highlighting the significantly altered microbial functional pathways in KEGG database between HPI and uninfected tissues. The differential metagenome functions were obtained using Metastats analysis with *q* value < 0.05. **e.** Significantly changed microbial carbohydrate-active enzymes families from the gastric microbiome of the HPI tissues using linear discriminant analysis (LDA) effect size (LEfSe) analysis (LDA>3). **f.** Significantly changed enzymes observed from the gastric microbiome of the HPI tissues compared to uninfected tissues from KEGG enzyme database. The differential metagenome enzymes were obtained using Metastats analysis with *q* value < 0.05. **g.** Only resistance gene hp1181, expressed in the *H. pylori*-infected tissues, was identified based on the comprehensive antibiotic research database (CARD). The differential metagenome gene was obtained using Metastats analysis with *q* value < 0.05.

**Suppl. Figure 2.**
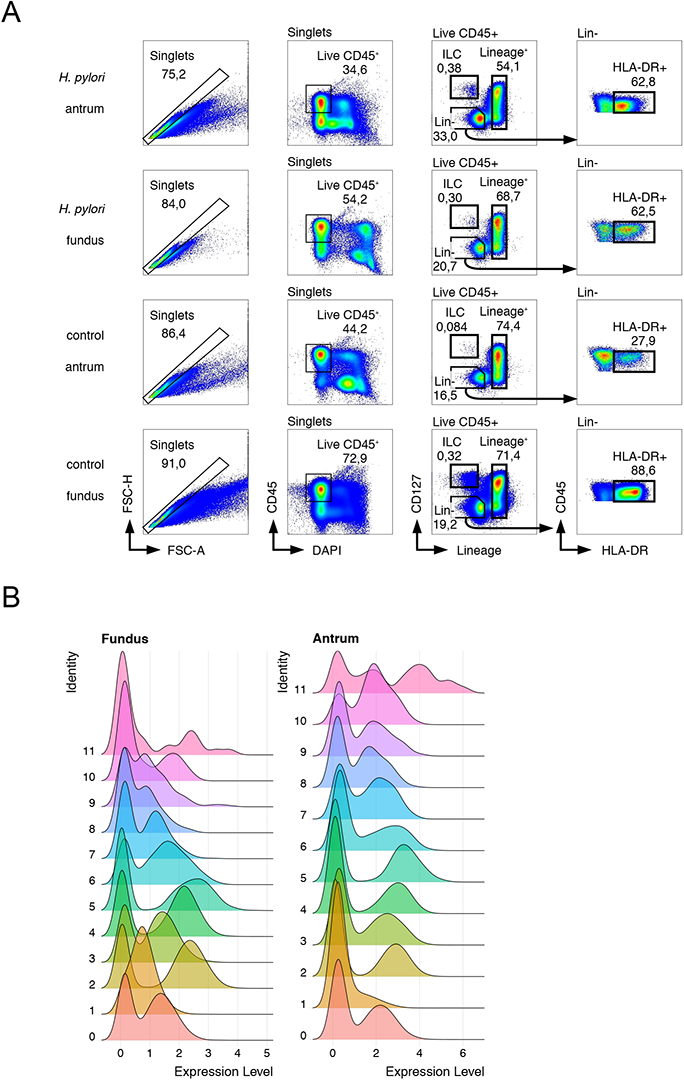
Sorting and clustering of immune cells from the stomach of HPI and uninfected tissues. **a.** Gating strategy for the sorting of selected immune cells from the stomach of HPI and uninfected tissues. **b.** Histograms showing expression of fundus-and antrum-associated hashtags within each cluster identified in Figure 2b.

**Suppl. Figure 3.**
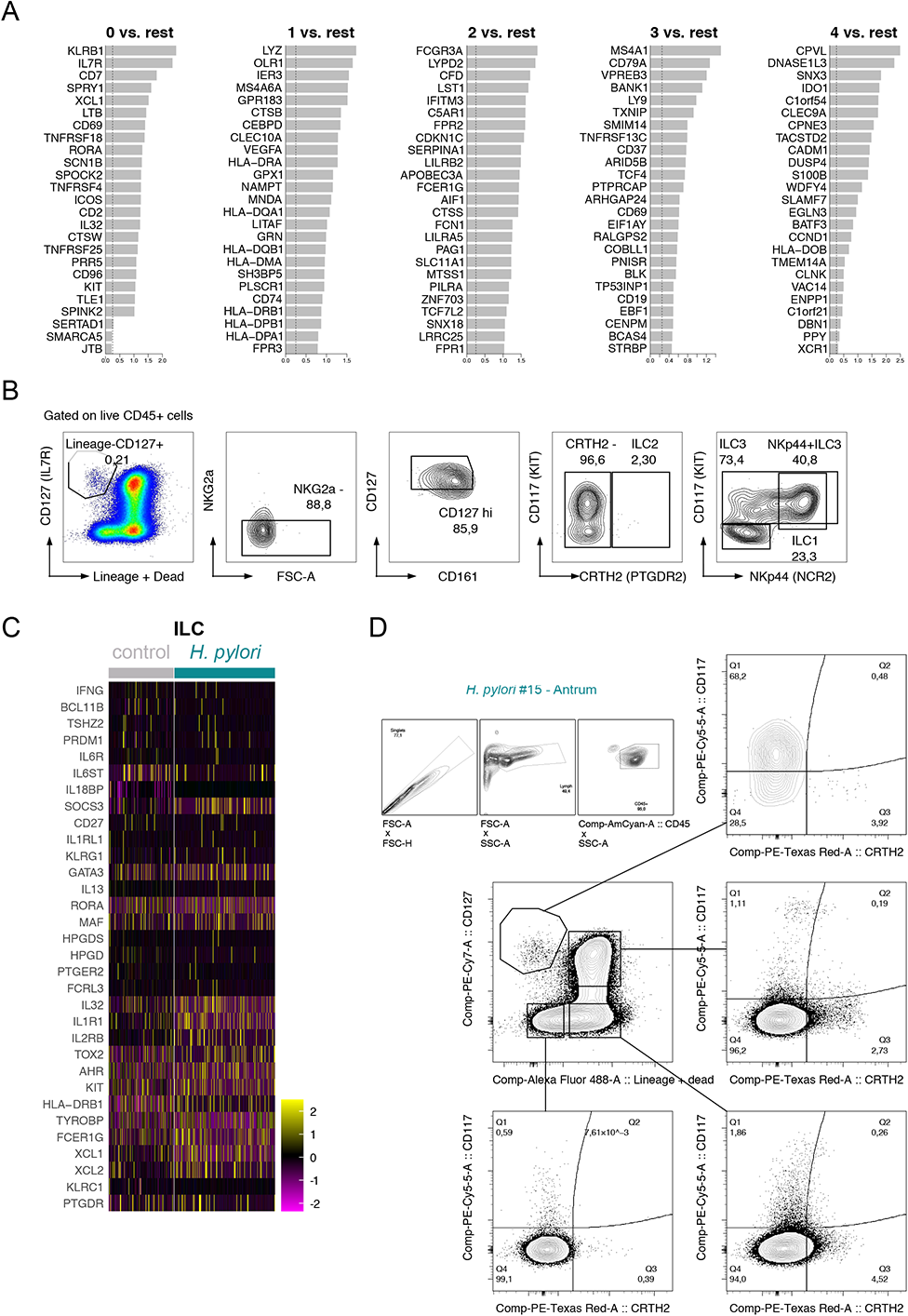
Innate immune cells from the HPI and uninfected tissues. **a.** Top differentially expressed genes in clusters shown in Figure 3a. **b.** Complete gating strategy used for the identification of ILC subsets in the gastric lamina propria. **c.** Transcriptomic insight of cells annotated as ILC in Figure 5a. **d.** Example of CD117 and CRTH2 protein expression in CD45^+^ cells other than Lineage^-^CD127^+^ ILCs.

**Suppl. Figure 4.**
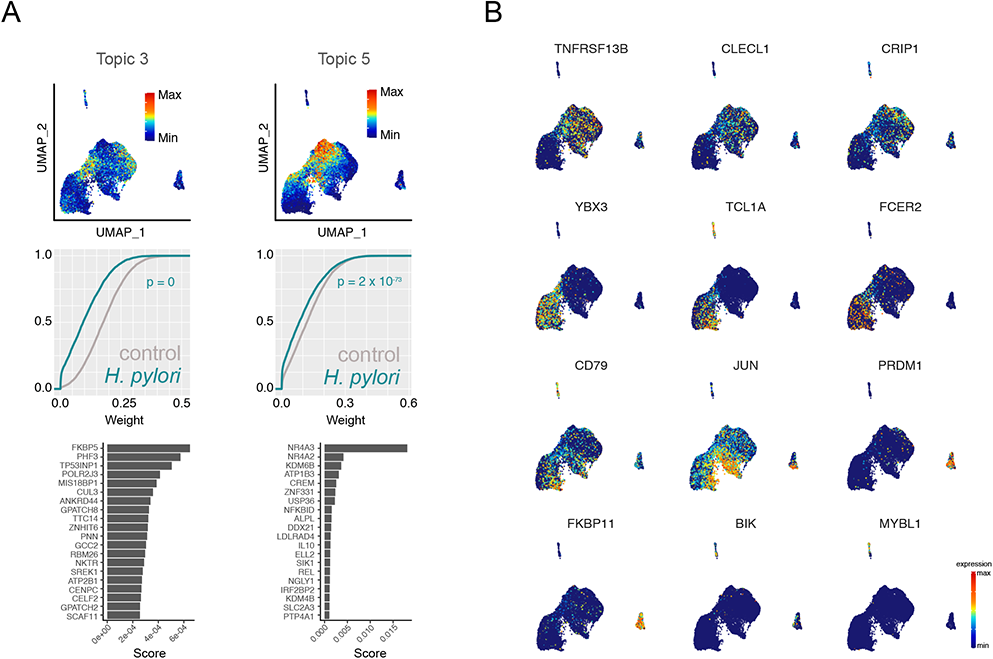
Additional B cell topic analysis in the HPI and uninfected tissues. **a**. B cell topics left out from analysis in Figure 4d. **b**. Weight of selected genes that define B cell topics.

**Suppl. Figure 5.**
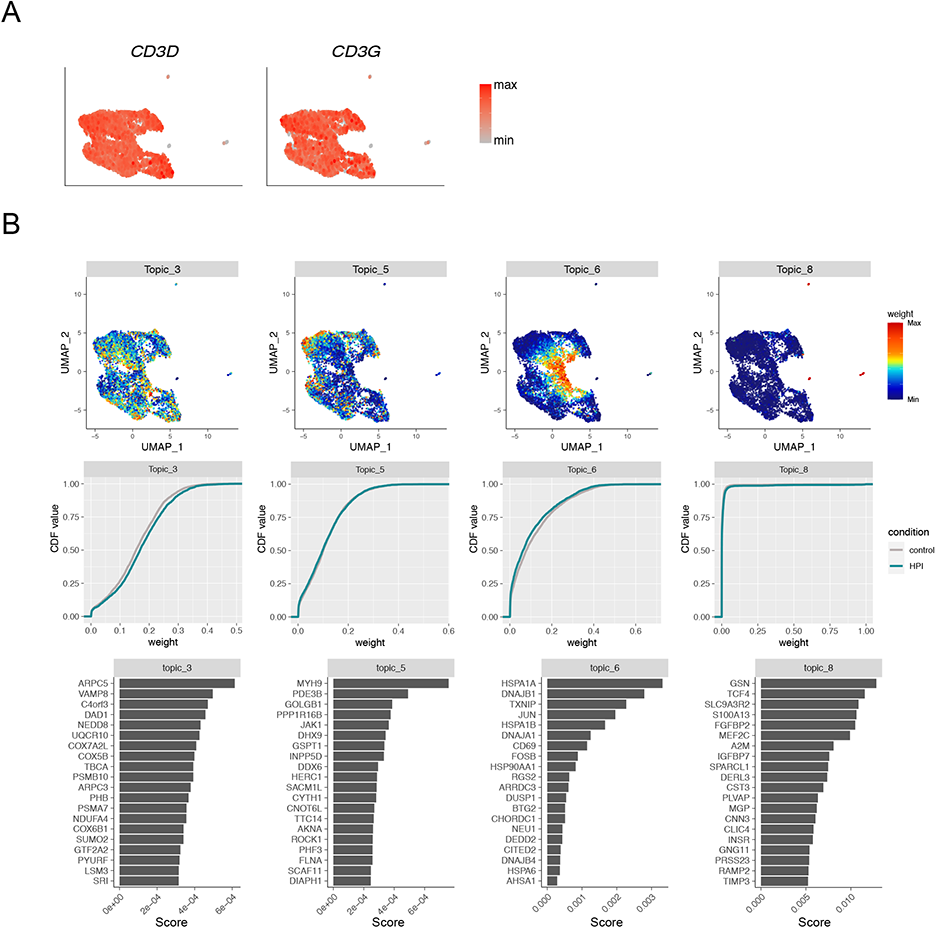
Additional T cell subset and topic analysis in the HPI and uninfected tissues. **a.** UMAP plots showing expression of *CD3D* and *CD3G* in T cells. **b.** T cell topics left out from analysis in Figure 5d.

**Suppl. Figure 6.**
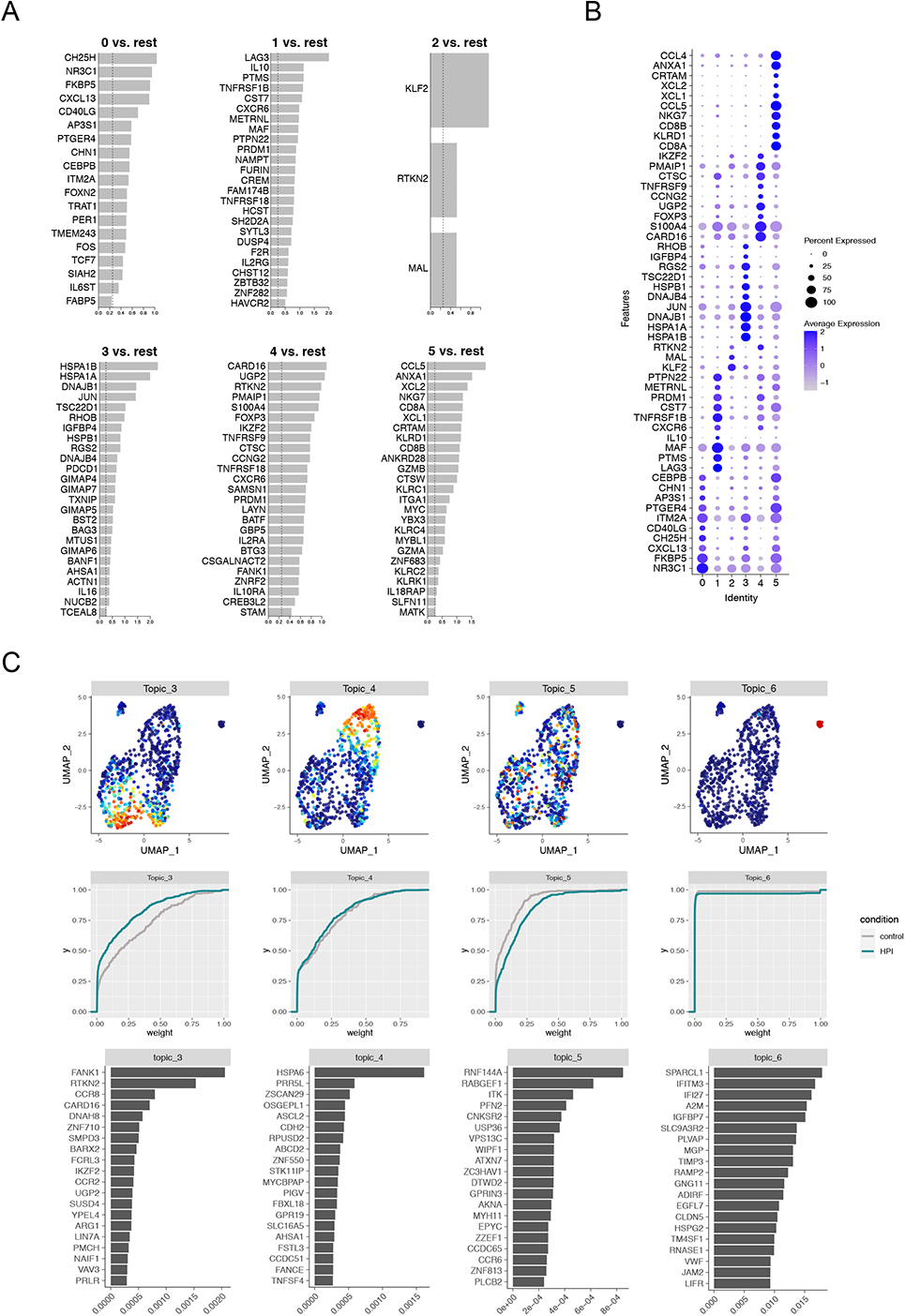
Characterization of activated CD4^+^ T cell subsets in the HPI and uninfected tissues. **a.** Top differentially expressed genes in clusters shown in Figure 6a. **b.** Dotplot showing expression of selected cell markers that were used to annotate clusters in Figure 6a. **c.** T cell topics left out from Figure 6d.

**Suppl. Figure 7.**
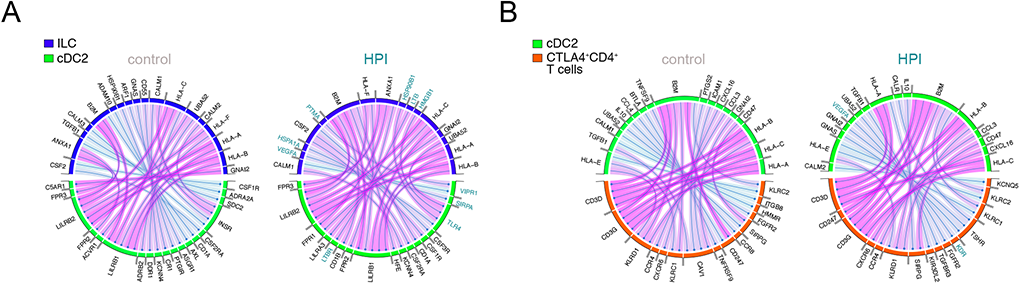
Innate and adaptive immune cells interact to form gastric TLS in the HPI tissues. **a**. Chord diagrams showing predicted communication pathways between ILC (ligands) and cDC2 (receptors) in HPI and uninfected tissues. **b**. Chord diagrams showing predicted communication pathways between cDC2 (ligands) and activated CD4^+^ T cells (receptors) in HPI and uninfected tissues.

## Notes

### Competing Interest Statement

E.J.V. has received research grants from F. Hoffmann-La Roche.

### Funding Statement

J.D. was supported by the Swedish Foundation for Strategic Research (SSF) (ICA16-0050), Svenska Lakaresallskapet (SLS-960584) and Karolinska Institutet. A.T. was supported by Erling-Persson Foundation (140604) and L. E. was supported by Soderbergs foundation.
E.J.V. was supported by grants from the Swedish Research Council, VR grant (K2015-68X-22765-01-6 and 2018-02533), Formas grant (FR-2016/0005), Cancerfonden (19 0395 Pj), and the Wallenberg Academy Fellow program (2019.0315). The computations and data handling were enabled by resources provided by the Swedish National Infrastructure for Computing (SNIC) at KTH partially funded by the Swedish Research Council through grant agreement no. 2018-05973. Ferring Phamaceuticals funded the centre where the sample collection and microbiome related experiments were carried out.

### Author Declarations

The study was approved by the Regional Ethical Board at Karolinska Institute, Stockholm, Sweden (Dnr: 2016/573-31)

